# Three-step rhythmic breathing exercise and COVID-19: A cross-sectional study

**DOI:** 10.1101/2021.07.07.21259527

**Authors:** Apurvakumar Pandya, Dileep Mavalankar, Pravin Maithia

## Abstract

**Introduction:** The present study assessed the prevalence of COVID-19 among people practicing three-step rhythmic breathing (3SRB) exercise and those who were not practicing any breathing exercises, including 3SRB exercise.

**Methods:** A community-based cross-sectional observational study was conducted. Data was collected using a self-constructed online google survey tool from July 2020 to August 2020.

**Results:** Out of a total 1083 sample, a higher proportion of the participants (41.3%) belonged to the 34-49 years age group, followed by the age group of 50-65 (32.5%). The sample was almost equally distributed; about 51.9% of the population was male, and 48.4% were female. The COVID-19 positivity was recorded almost double (3.1%) in groups not practicing 3SRB exercises compared to a group (1.3%) practicing 3SRB exercises. Furthermore, the practice of 3SRB was significantly associated with a lower percentage of COVID-19 infection (p=0.046).

**Conclusions:** Practice of 3SRB is significantly associated with a lower percentage of COVID-19 infection. A future study with a robust methodology is warranted to validate the findings of this study and determine the effects of 3SRB on physiological and biological markers.

## Introduction

The coronavirus disease 2019 (COVID-19) pandemic has become a significant cause of physical & psychological distress and deaths worldwide. Many European and Asian countries have faced a second wave of the pandemic while India is experiencing the second wave at present The COVID-19 damages the respiratory tract, obstructing airflow.^1^ It triggers asthma attacks and causes acute respiratory distress syndrome (ARDS).^2^ Deep breathing exercises that clear the lungs and strengthen lung function may be especially beneficial for people with these conditions.^3,4^ Evidence indicates that breathing exercises improve lung functions,^5,6^ boost immunity,^7^ and manages stress.^8^ However, the effect of breathing exercise on COVID-19 infection is not yet established.

Three-Step Breathing exercise is a breathing pattern that aids an individual ‘breathe in rhythm with nature. It is a slow and deep breathing technique founded by Yogi Sri Soli Tavaria. It is also known as refining exercises and is believed to cleanse impurities from the mind and the body. Studies on the effect of Three Step Rhythmic Breathing (3SRB) on COVID-19 remain unknown. Anecdotal experience that breathing exercises can strengthen the lungs and may be beneficial for reducing the impact of COVID-19 before, during, and after it strikes. The present study attempted to determine the prevalence of COVID-19 among people practicing 3SRB exercise and those who are not practicing 3SRB exercise in India.

## Methods

A community-based cross-sectional observational study was conducted. Data was collected using a self-constructed online google survey tool from July 2020 to August 2020. The data entry was done using Microsoft Office Excel 2019. Descriptive statistics of frequency, percentages, mean and standard deviation (SD) were used to summarize the data. Categorical variables are represented as frequencies and percentages and analyzed using the χ 2 test or Fisher’s exact test. All statistical analyses were performed using SPSS (Statistical Package for the Social Sciences) version 23.0 software (SPSS, Inc). A 2-sided α (Alpha) level at less than 0.05 or 0.01 was considered statistically significant.

## Results

Table 1 shows the socio-demographic characteristics of the study respondents. A higher proportion (40.3% for group A and 42.4% in group B, respectively) of the participants belonged to the 34-49 years age group, followed by the age group of 50-65 years (31.4% and 32.5% respectively). The mean age in group A was 45.5±13.6 and 47.5±14.5 in group B. About 51.9% of the population was male, and 48.4% were female.

**Table 1.**
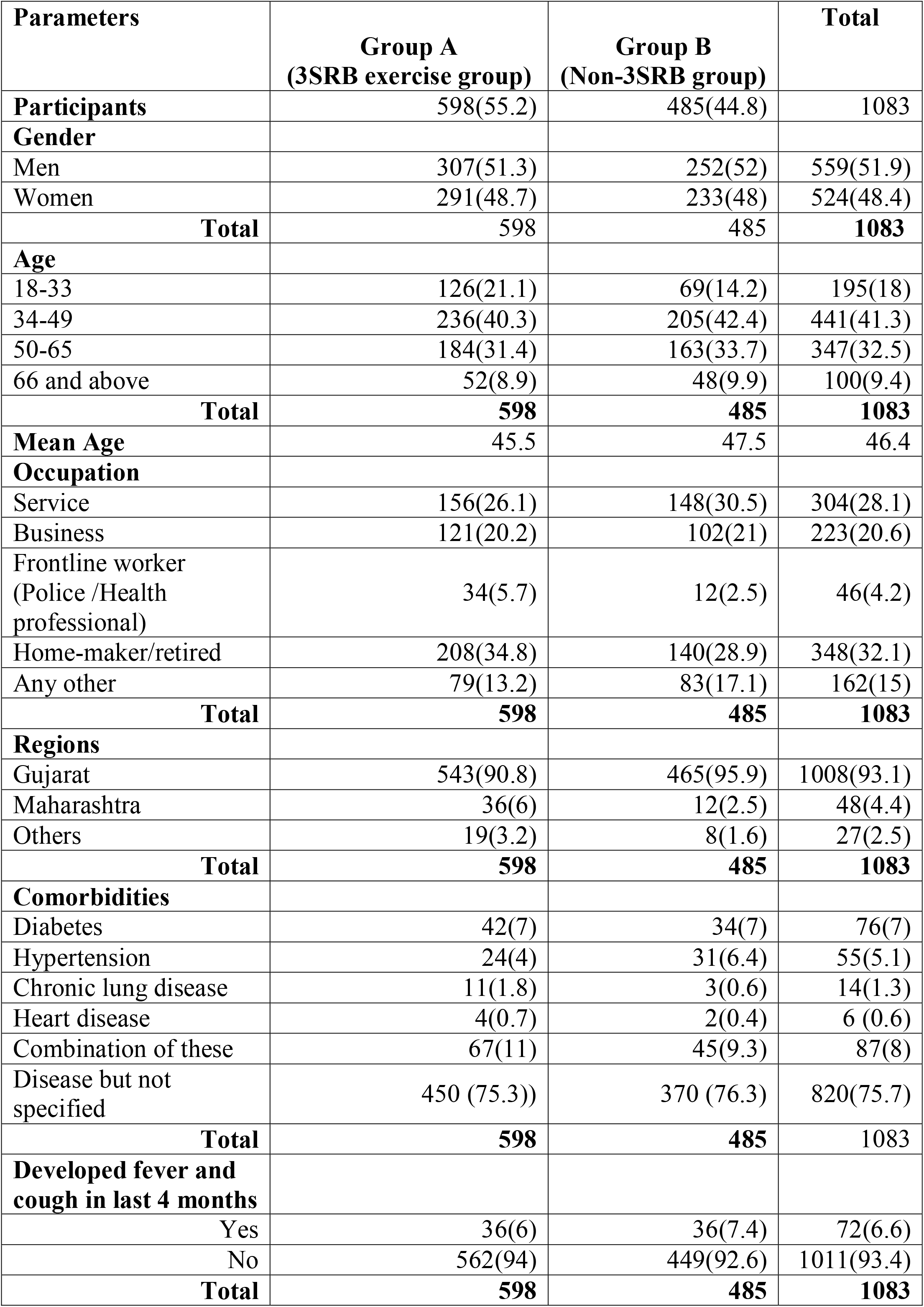
Demographic profile of study participants.

About 32.1% of participants were homemakers or retired, 28.1% were doing service, and 20.6% were engaged in business. In the study, the majority of respondents (93.1%) were from Gujarat, followed by Maharashtra (4.4%), and the rest were from other States (2.5%). Current comorbidities included diabetes (7%), hypertension (5.1%), followed by the highest prevalence of a comorbid condition of diabetes, hypertension, or heart disease (8%). Table 1 presents the demographic characteristics of study participants.

### Characteristics of persons practicing 3SRB

Total 598 participants were practicing 3SRB. About 47.9% were practicing for less than a year, and 79.8% were practicing daily. Table 2 presents the characteristics of 3SRB practice. Out of the total, 11.7% of 3SRB practitioners were reported treating other chronic diseases. These diseases were diabetes (2.3%), Arthritis /Spondylitis (2%), Blood pressure (1.8%), Psychological distress such as depression, anxiety and stress (1.8 %), Asthma (1.2%), hypertension & thyroid (1% respectively), thyroid (1%), and cardiovascular disease (0.2%). Many participants (1.7%) reported treating Parkinson‘s, Psoriasis, etc.

**Table 2.** Characteristics of Refining Exercise Group.

| **Experience of Refining exercise** | ***f*** | **%** |
| --- | --- | --- |
| <1 year | 244 | 47.9 |
| 1 to 5 yrs | 176 | 29.4 |
| 6 to 10 yrs | 57 | 9.5 |
| >10 yrs | 121 | 20.2 |
| Total | 598 | 100 |
| **Frequency (days in week)** |  |  |
| 1 to 2 | 12 | 2.0 |
| 3 to 4 | 34 | 5.7 |
| 5 to 6 | 93 | 15.6 |
| Everyday | 425 | 76.8 |
| Total | 598 | 100 |
| **Duration of refining exercise** |  |  |
| 5 minutes | 37 | 6.2 |
| 6 to 15 minutes | 135 | 22.6 |
| 16-30 minutes | 246 | 41.1 |
| More than 30 minutes | 180 | 30.1 |
| Total | 598 | 100 |
| **Treating chronic diseases** |  |  |
| Yes | 70 | 11.7 |
| No | 528 | 88.3 |
| Total | 598 | 100 |

### Prevalence of COVID-19 in the study population

Overall prevalence was 2.1% in the study population; 1.3% in group A followed by 3.1% in group B, as shown in Table 3. The association between practice and non-practitioners of 3SRB and COVID-19 was significant (p=0.046).

**Table 3.**
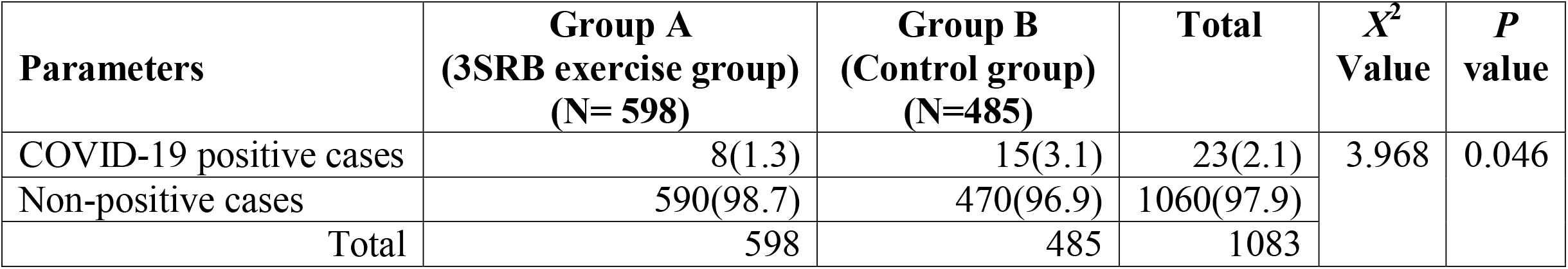
COVID-19 positive cases in group A & B.

| **Parameters** | **Group A**<br>**(3SRB exercise group)**<br>**(N= 598)** | **Group B**<br>**(Control group)**<br>**(N=485)** | **Total** | **$X^2$**<br>**Value** | **$P$**<br>**value** |
| --- | --- | --- | --- | --- | --- |
| COVID-19 positive cases | 8(1.3) | 15(3.1) | 23(2.1) | 3.968 | 0.046 |
| Non-positive cases | 590(98.7) | 470(96.9) | 1060(97.9) |  |  |
| Total | 598 | 485 | 1083 |  |  |

Table 4 presents data on COVID-19 positive cases in the family and the residential areas among both groups. In the total sample, 4.1% of participants’ family members were tested positive for COVID-19 and was a non-significant association for COVID-19 infection across two groups (p=0.827). Nearly half of participants (48.6%) had COVID-19 cases in their residential area, whereas 33.7% reported COVID-19 cases in their wards. COVID-19 cases in their resident area and ward were not statistically significant with COVID-19 (p=0.076).

**Table 4.** COVID-19 positive cases in the family and in the residential areas.

| **Parameters** | **Group A** | **Group B** | **Total** | **$X^2$**<br>**value** | ***P***<br>**value** |
| --- | --- | --- | --- | --- | --- |
| Family members tested COVID-19 positive | 25(4.2) | 19(3.9) | 44(4.1) | 0.0476 | 0.827 |
| Family members not COVID-19 positive | 573(95.8) | 466(96.1) | 1039(95.9) |  |  |
| **Total** | **598** | **485** | **1083** |  |  |
| COVID cases in ward | 189(31.6) | 176(36.5) | 365(33.7) | 5.153 | 0.076 |
| COVID cases in resident area | 309(51.7) | 217(44.7) | 526(48.6) |  |  |
| No COVID cases in our ward/area | 100(16.7) | 92(19) | 192(17.7) |  |  |
| **Total** | **598** | **485** | 1083 |  |  |

### Association between independent variables and COVID-19 positivity across Two Groups

Table 5 presents an association between independent variables with COVID-19 in both groups. More than half (65.2%) positive cases belonged to the 34-49 years of age group, and most men (87%) were affected. In terms of occupation, persons from the business (56.5%) and service (34.8) category were primarily affected with COVID-19.

**Table 5.** Association between independent variables and COVID-19 positivity across groups.

| **Parameters** | **Group A**<br>**(n=8)** | **Group B**<br>**(n=15)** | **Total**<br>**(N=23)** |
| --- | --- | --- | --- |
| **Age** |  |  |  |
| 18-33 | 1(12.5) | 2(13.3) | 3(13) |
| 34-49 | 6(75) | 9(60) | 15(65.2) |
| 50-65 | 1(12.5) | 4(26.7) | 5(21.7) |
| **Total** | **8** | **15** | **23** |
| **Mean age** |  |  |  |
| **Gender** |  |  |  |
| Male | 7(87.5) | 13(86.7) | 20(87) |
| Female | 1(12.5) | 2(13.3) | 3(13) |
| **Total** | **8** | **15** | **23** |
| **Occupation** |  |  |  |
| Service | 4(50) | 4(26.7) | 8(34.8) |
| Business | 4(50) | 9(60) | 13(56.5) |
| Frontline workers/COVID-19 Warriors | 0 | 1(6.7) | 1(4.3) |
| Home-maker/retired | 0 | 1(6.7) | 1(4.3) |
| **Total** | **8** | **15** | **23** |
| **Comorbidities among COVID positive** |  |  |  |
| Hypertension | 2(25) | 7(46.7) | 9(39.1) |
| Combination of diabetes/hypertension | 6(75) | 8(53.3) | 14(60.9) |
| **Total** | **8** | **15** | **23** |
| **Treatment** |  |  |  |
| Hospitalization | 3(37.5) | 5(33.3) | 8(34.8) |
| Home treatment | 5(62.5) | 10(66.7) | 15(65.2) |
| **Total** | **8** | **15** | **23** |
| **Family members tested COVID positive** |  |  |  |
| Yes | 1(12.5) | 6(40) | 17(30.4) |
| No | 7(87.5) | 9(60) | 16(69.6) |
| **Total** | **8** | **15** | **23** |
| **COVID-19 cases in residential areas** |  |  |  |
| COVID cases in resident area | 7(87.5) | 14(93.3) | 21(91.3) |
| COVID cases in the ward | 1(12.5) | 1(6.7) | 2(8.7) |
| **Total** | **8** | **15** | **23** |

As shown in Table 6, symptoms of fever and cough were present in 47.8% of positive cases, which means the presence of both symptomatic and asymptomatic cases. Symptoms were more frequent in positive individuals (47.8%) than those not diagnosed with COVID-19 (6.1%).

**Table 6.** Symptoms of fever and cough in last 3 months.

| **Parameters** | **Group A**<br>**(3SRB exercise group)**<br>**(N= 598)** | **Group B**<br>**(Control group)**<br>**(N=485)** | **Total** |
| --- | --- | --- | --- |
| COVID-19 positive cases* | 4 (50) | 7 (46.7) | 11 (47.8) |
| Non-positive cases** | 35(5.9) | 30 (6.4) | 65 (6.1) |
| Total | 39 (6.5) | 37 (7.6) | 67 (7) |

\*(n=8 for Group A; n=15 for Group B) \*\*(n=590 for Group A; n=470 for Group B)

About 43.5% of COVID-19 positive cases were from Surat, followed by 26.1% from Ahmedabad. Both of these cities were marked with a high case of COVID-19 in the State during the study period. Table 7 presents the distribution of COVID-19 positive cases across cities.

**Table 7.**
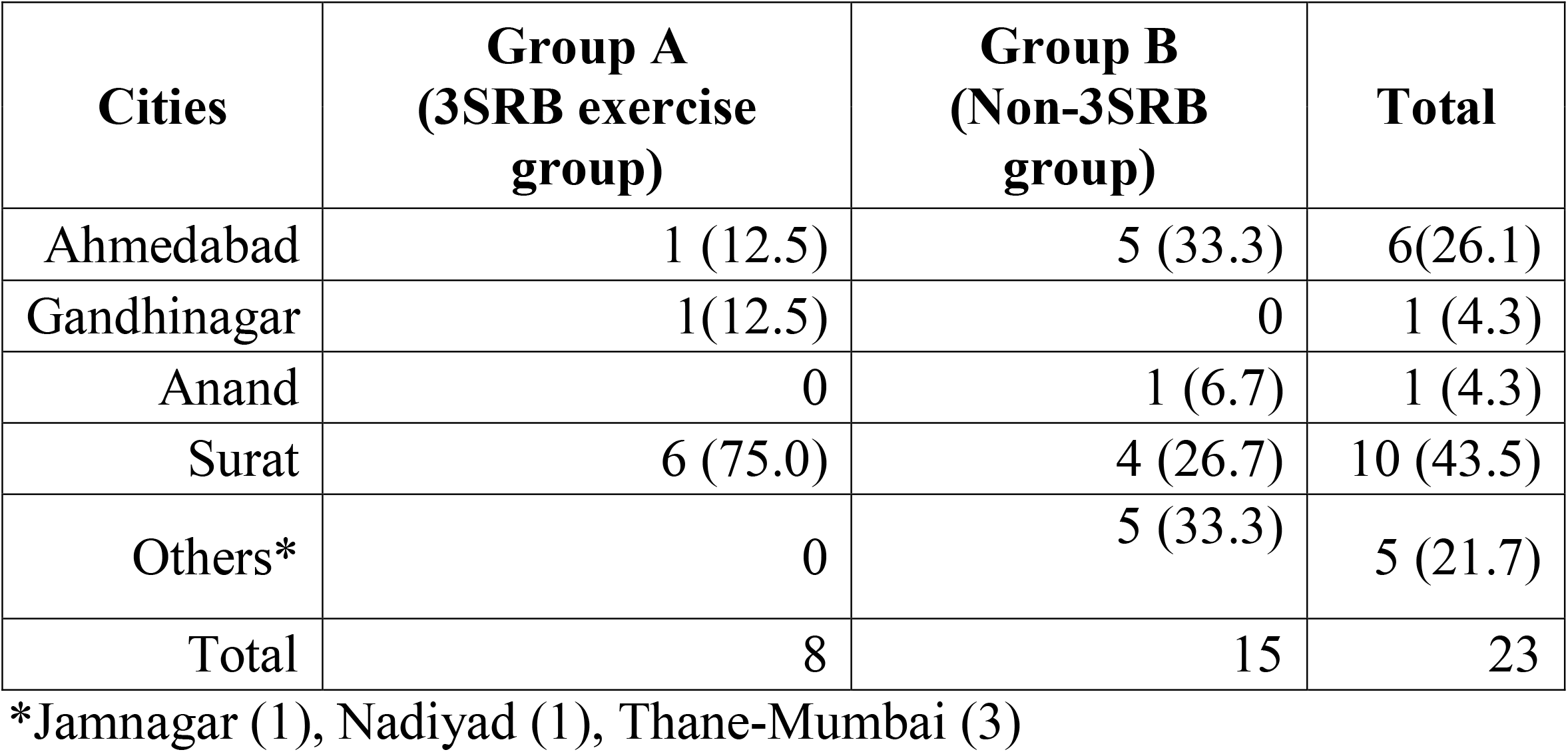
Distribution of COVID-19 prevalence across cities.

| **Cities** | **Group A**<br>**(3SRB exercise**<br>**group)** | **Group B**<br>**(Non-3SRB**<br>**group)** | **Total** |
| --- | --- | --- | --- |
| Ahmedabad | 1 (12.5) | 5 (33.3) | 6(26.1) |
| Gandhinagar | 1(12.5) | 0 | 1 (4.3) |
| Anand | 0 | 1 (6.7) | 1 (4.3) |
| Surat | 6 (75.0) | 4 (26.7) | 10 (43.5) |
| Others* | 0 | 5 (33.3) | 5 (21.7) |
| Total | 8 | 15 | 23 |

\*Jamnagar (1), Nadiyad (1), Thane-Mumbai (3)

As presented in Table 8, about 56.6% of the study participants infected with COVID-19 were practicing yoga, meditation, exercise, or any combination of these, and more than half of them (65.2%) have reported using herbs for recovery.

**Table 8.** Association between other exercises, use of herbs and COVID-19.

| **Parameters** | **Group A**<br>**(3SRB exercise group)** | **Group B**<br>**(Non-3SRB group)** | **Total**<br>**(N=23)** |
| --- | --- | --- | --- |
| **Other exercises** |  |  |  |
| Yoga/meditation or exercise | 5 (62.5) | 8 (53.3) | 13 (56.5) |
| None | 3(37.5) | 7(46.7) | 10 (43.5) |
| Total | **8** | **15** | 23 (100) |
| **Use of herbs** |  |  |  |
| Yes | 4 (50.0) | 11 (73.3) | 15 (65.2) |
| No | 4 (50.0) | 4 (26.7) | 8 (34.8) |
| Total | **8** | **15** | **23** |

## Discussion

The study disclosed the prevalence of COVID-19 as 2.1% in the entire study population, corresponding to 23 individuals infected with the novel coronavirus. About 1.3% positivity was observed in group A who were practicing 3SRB or refining exercise, and 3.1% prevalence in group B, who were not practicing refining exercise. The prevalence is higher than the recently conducted study among Healthcare workers (1.8%)^11^ and the study conducted in Brazil, which reported a prevalence of 2.1% COVID-19 infection.^12^ Further, an association between practice and non-practitioners of 3SRB and COVID-19 was significant (p=0.046).

The majority of study participants infected with COVID-19 were engaged in business or service, suggesting occupation as a critical determinant of COVID-19. As such, physical activity and or exercise were beneficial in ameliorating some of the respiratory difficulties and psychological distress;^13,14^ which is endorsed by the present study‘s findings. However, due to small numbers of positive cases, a significant effect of exercise could not be determined.

Positivity in a family, ward, or residential area was associated with COVID-19 infection; however, the association was not significant. In the total sample, 4.1% of respondents’ family members were tested positive for COVID-19. A positive case in the family is an expected result in a fast-evolving epidemic like COVID-19. ^15, 16^ Nevertheless, at this point, it is possible to observe a higher probability of positive contacts when the family member is detected COVID-19 positive. Recent systematic reviews conducted by Shah et al.^17^indicated that the secondary attack rate in household contacts was found to be ranging between 4.6 and 49.56%. The study did not evaluate contributing factors of COVID-19 positivity and factors such as age, duration of the positive case in the family, house dimensions, and the number of rooms in the family. Recent studies indicate that spouses and the elderly are more prone to secondary infection than the younger population (supported through reported odds). Minors (<18/<20 years) are at lower risk of SAR.^18,19^

Among all positive individuals, 52.2% were asymptomatic, and the proportion was lower than other studies that reveal the frequency of symptoms as high as 81%.^16^ Moreover, symptoms were more frequent in positive individuals (47.8% compared to 6.1% in negative individuals), indicating validity for the test and is in concordance with results from other studies.^17, 18^ The information about symptoms was referent to their occurrence in the last three months in this study. The longer lag time between symptoms occurrence and reporting could increase their frequencies, but it increases the possibility of memory bias.

### Strength and limitations of the study

To the best of our knowledge, the study is the first to assess the prevalence of COVID-19 infection among people practicing 3SRB and those who were not practicing. Our study has some notable limitations. First, we compared individuals practicing the 3SRB exercise with non-practicing individuals. The data collection was facilitated through volunteers of 3SRB exercise groups, which may introduce participant selection bias. Therefore, our results should be interpreted carefully. Second, we have used self-reported COVID-19 positivity, and not performed COVID-19 tests may present self-reporting bias. This may have generated a socially desirable response, and many might have intentionally hidden their status of COVID-19 infection due to prevalence stigma. Third, the 3SRB exercise emerged as beneficial in protecting them from COVID-19; however, the findings can not be easily attributable only to 3SRB as confounding variables such as yoga, meditation, pranayama, and herbs might have influenced the outcomes. Finally, a cross-sectional study design cannot establish causality between various socio-demographic factors and COVID-19.

## Conclusion

The current study indicated that the prevalence of COVID-19 is lower among people practicing 3SRB compared to those who are not practicing 3SRB and the difference was statistically significant. The 3SRB exercise can be a promising practice for the prevention and may promote better recovery from the infection. Prospective large-scale studies should consider controlling confounding variables. A future study using a robust methodology such as a randomized control trial or case-control trial should be conducted to validate the findings of this study and determine the effects of 3SRB on COVID-19 infection, disease progression, and management using biological markers.

## Data Availability

Data will be available upon request

## Declaration

## Acknowledgment

Authors appreciate participants for their active participation in the study.

## Ethics approval

This study was performed in line with the principles of the Declaration of Helsinki. Informed consent was obtained from all individual participants included in the study.

